# Trust in scientists and conspiracy beliefs predict online misinformation susceptibility and fake news detection: a cross-sectional study in Greece

**DOI:** 10.1101/2025.11.18.25340464

**Authors:** Aglaia Katsiroumpa, Ioannis Moisoglou, Olympia Konstantakopoulou, Olga Galani, Maria Tsiachri, Petros Galanis

## Abstract

**Background:** Online misinformation has grown significantly with the widespread use of the internet and the ease of sharing content through social media, often without rigorous fact-checking or scientific validation. In this context, trust in scientists and conspiracy beliefs may affect online misinformation susceptibility and fake news detection.

**Objective:** To examine the effect of trust in scientists and conspiracy beliefs on online misinformation susceptibility and fake news detection. Additionally, we investigated the association between demographic variables and online misinformation susceptibility and fake news detection.

**Methods:** A cross-sectional study was carried out in Greece, with data collected through an online survey in August 2025. Trust in scientists was assessed using the Trust in Scientists Scale. Conspiracy beliefs were measured using the Conspiracy Mentality Questionnaire. Participants’ susceptibility to online misinformation was evaluated using the Online Misinformation Susceptibility Scale. Their ability to detect online fake news was assessed through the fake news detection scale.

**Results:** Our multivariable analysis identified that lower trust in scientists is associated with higher online misinformation susceptibility. Participants who believed in conspiracy behaviors showed also higher levels of misinformation susceptibility. Moreover, lower financial status and lower interest in politics were associated with misinformation susceptibility. We found a positive association between trust in websites and misinformation susceptibility. Also, we found a negative association between age and misinformation susceptibility. Our multivariable linear regression model showed that participants who believe in conspiracy beliefs had a lower ability to detect fake news. Moreover, we found a positive association between trust in sciences and fake news detection. Finally, we found a negative association between interest in politics and fake news detection.

**Conclusion:** Our findings showed associations between trust in scientists, conspiracy beliefs, online misinformation susceptibility and fake news detection. Moreover, several demographic variables were associated with misinformation susceptibility and fake news detection. Identification of predictors of misinformation susceptibility and fake news detection is crucial to define high-risk groups and develop appropriate interventions to confront these issues.

## Introduction

In the digital age, the proliferation of misinformation and fake news has emerged as a significant challenge to public discourse, democratic processes, and individual decision-making. Misinformation refers to false or misleading information that is shared without the intent to deceive (Hameleers & Brosius, 2022), whereas fake news constitutes deliberately fabricated content designed to mimic legitimate news sources, often with the aim of manipulating public opinion or generating profit (Lazer et al., 2018). Misinformation, whether disseminated unintentionally or with deliberate intent, refers to inaccurate or misleading information that can have significant adverse consequences for both individuals and society (Borukhson et al., 2022; Ecker et al., 2020). Psychological research shows that repetition reliably increases perceived truth (the illusory truth effect), helping explain why repeated claims online can appear credible even when false; this effect extends to headlines and conspiratorial content and is partly driven by processing fluency (Udry & Barber, 2024). At the same time, evidence from cognitive and judgment–decision-making studies indicates that analytic/deliberative processing improves truth discernment for headlines and full-length articles, countering intuitive acceptance of false claims and supporting dual-process accounts of misinformation resistance (Bago et al., 2020).

The rise of digital communication platforms has transformed the production, dissemination, and consumption of information. Among the most pressing challenges in this landscape is the spread of online misinformation, defined as false, inaccurate, or misleading information shared via digital media and especially social media, often without malicious intent. In contrast, disinformation is deliberately crafted to deceive while mimicking legitimate news formats and genres. The digital environment—characterized by speed, scale, and limited editorial gatekeeping—enables rapid amplification of such content across social media, blogs, forums, and encrypted messaging applications (Aïmeur et al., 2023; Pennycook & Rand, 2021; Udry & Barber, 2024). Online misinformation manifests in multiple forms, including fabricated news articles, manipulated images and videos, pseudoscientific claims, and misleading statistics (Del Vicario et al., 2016).

Understanding the predictors of misinformation susceptibility is essential for developing effective interventions and enhancing public resilience against false or misleading information. A growing body of interdisciplinary research has identified a range of psychological and socio-demographic factors that influence individuals’ vulnerability to misinformation and fake news.

In particular, cognitive traits such as analytical thinking, cognitive reflection, and numeracy are consistently associated with lower susceptibility to misinformation. Individuals with stronger analytical reasoning skills are better able to discern between true and false information, even when content aligns with their ideological beliefs (Roozenbeek et al., 2022; Sultan et al., 2024). Conversely, myside bias—the tendency to evaluate information based on its alignment with one’s own views—has been shown to increase susceptibility, particularly in politically polarized contexts (Hubeny et al., 2025). Moreover, recent studies have identified several personality traits that predict misinformation acceptance. For example, low openness to experience, high grandiose narcissism, and low emotional stability are linked to higher belief in false information (Piksa et al., 2022). Additionally, dispositional optimism and anxiety may influence how individuals engage with and interpret misleading content (Kaufman et al., 2025).

Demographic factors such as age, education, and political identity also play a role. A recent meta-analysis suggests that older adults may be more skeptical and cautious in their judgments, though they also engage more frequently with misinformation online (Sultan et al., 2024). Educational attainment does not consistently predict susceptibility, challenging assumptions that higher education always confers protection against misinformation (Scherer & Pennycook, 2020). Political identity, particularly partisan alignment, influences how individuals evaluate news content, with ideological congruency increasing belief in misinformation (Kim et al., 2022). On the other hand, individuals who identified themselves as democrats have higher discrimination ability between fake and true news (Sultan et al., 2024). Heavy reliance on social media for news consumption is consistently associated with increased belief in misinformation. This relationship is mediated by exposure to unverified content, algorithmic amplification, and the absence of editorial gatekeeping. Emotional engagement and rapid sharing behaviors further exacerbate the spread of false information (Ali Adeeb & Mirhoseini, 2023).

Distrust in science has emerged as a critical barrier to effective public communication, policy implementation, and societal resilience in the face of global challenges. This phenomenon is associated with a range of adverse outcomes across domains such as public health, information integrity, institutional legitimacy, and social cohesion. For instance, during the COVID-19 pandemic, low trust in scientific authorities was strongly associated with vaccine hesitancy, refusal to adhere to public health guidelines, and increased reliance on unverified treatments. In particular, during the COVID-19 pandemic, individuals who lacked confidence in scientific sources were more likely to seek information from social media and alternative outlets, which often lacked editorial oversight and promoted misleading or conspiratorial content. A two-wave panel study in the U.S. found that distrust in science directly increased reliance on social media for pandemic-related information, which in turn heightened susceptibility to misperceptions about COVID-19 (Lee et al., 2024). Additionally, low trust in scientific authorities contributed to non-compliance with public health recommendations, including mask-wearing, social distancing, and vaccination. Individuals skeptical of science were significantly less likely to support social distancing measures, even during the early stages of the pandemic when public consensus was relatively high (Hatton et al., 2022). Individuals who distrust scientific institutions are more likely to believe and disseminate misinformation and conspiracy theories, particularly when such content is framed in pseudo-scientific language. Experimental studies have shown that high trust in science can paradoxically increase vulnerability to scientifically styled misinformation, unless accompanied by critical evaluation skills (O’Brien et al., 2021). Moreover, distrust in science often leads individuals to seek information from alternative sources, such as social media or partisan outlets, which may lack credibility and promote false narratives (Erisen & Erisen, 2024).

Conspiracy theories—explanatory narratives that attribute significant events to secret plots by powerful actors—have become increasingly prevalent in digital and political discourse (Freeman & Bentall, 2017; Uscinski et al., 2022). While often dismissed as fringe beliefs, a growing body of research demonstrates that conspiracy theories can have serious psychological, social, and political consequences for individuals and society.

Belief in conspiracy theories is associated with feelings of powerlessness, uncertainty, and mistrust. These beliefs can erode individuals’ sense of agency and reduce their engagement with civic and health-related behaviors. For example, exposure to conspiracy narratives about climate change or vaccination has been shown to decrease intentions to act pro-environmentally or to vaccinate oneself or one’s children, even when the information is demonstrably false (Butter & Knight, 2020; Jolley & Douglas, 2014). Conspiracy beliefs also correlate with science denialism, political apathy, and support for non-normative political actions, including protest and civil disobedience. These effects are particularly pronounced when conspiracy theories frame institutions as corrupt or malevolent, leading individuals to disengage from democratic processes (Butter & Knight, 2020). At the interpersonal level, conspiracy beliefs can strain relationships and reduce social trust. Studies have found that individuals in relationships with conspiracy believers report lower relationship satisfaction, increased social isolation, and mistrust of close others. These effects may stem from the way conspiracy theories encourage suspicion and reduce empathy, framing others as deceptive or complicit in hidden agendas (Forgas, 2025; Green et al., 2023; Toribio-Flórez et al., 2023). Moreover, publicly expressing conspiracy beliefs can damage one’s reputation and perceived credibility. Individuals who share anti-science or politically charged conspiracy theories are often judged as less trustworthy or competent, particularly in fields where evidence-based reasoning is valued (Forgas, 2025).

In this context, identifying the predictors of susceptibility to online misinformation and the ability to detect fake news is crucial for understanding the underlying mechanisms and developing appropriate interventions to reduce misinformation and vulnerability to fake news. However, to the best of our knowledge, there are no studies that investigate the impact of trust in scientists and conspiracy beliefs on online misinformation susceptibility and fake news detection. Thus, our study examined trust in scientists and conspiracy beliefs as potential predictors of susceptibility to online misinformation and the ability to detect fake news. In addition, several demographic variables were investigated to assess their predictive value regarding misinformation vulnerability and fake news detection

## Methods

### Study design

A cross-sectional study was carried out in Greece, with data collected through an online survey in August 2025. The questionnaire was created using Google Forms and distributed via Facebook, Instagram, and LinkedIn, resulting in a convenience sample. Participants were eligible if they met the following criteria: (1) were adults, (2) spent at least 30 minutes per day on the internet or social media, and (3) gave informed consent to participate. The study followed the Strengthening the Reporting of Observational Studies in Epidemiology (STROBE) guidelines (Von Elm et al., 2008). Sample size was calculated using G*Power v.3.1.9.2. With nine predictors included in the multivariable models, an anticipated effect size of 0.04 for each predictor-outcome relationship, a statistical power of 95%, and a 5% margin of error, the required sample size was estimated at 327 participants.

### Measurements

We collected data on various demographic characteristics of the participants, including gender (male or female), age (treated as a continuous variable), and educational attainment (elementary school, middle school, high school, university degree, or MSc/PhD). Financial status was assessed using a self-rated scale ranging from 0 (very poor) to 10 (excellent). Participants also rated their trust in websites to accurately report news on a scale from 0 (not at all) to 10 (completely), their interest in politics on a similar 0 to 10 scale, and reported the number of hours spent daily on the internet or social media (continuous variable).

To evaluate participants’ trust in scientists, we utilized the Trust in Scientists Scale (TISS), which comprises 12 items assessing four primary dimensions: integrity, competence, benevolence, and openness (Cologna et al., 2025). Due to the high correlation among these dimensions, the integrity subscale was selected as a representative measure for our study. This subscale includes three items: “How honest or dishonest are most scientists?”, “How ethical or unethical are most scientists?”, and “How sincere or insincere are most scientists?”. Participants responded using a five-point Likert scale, ranging from 1 (very dishonest/unethical/insincere) to 5 (very honest/ethical/sincere). The integrity score was calculated by averaging the responses to these three items, resulting in a final score between 1 and 5, with higher scores indicating stronger trust in scientists. The validated Greek version of the scale was used (Cologna et al., 2025). In our sample, the integrity subscale demonstrated strong internal consistency, with a Cronbach’s alpha of 0.908.

To assess participants’ general conspiracy beliefs, we administered the Conspiracy Mentality Questionnaire (CMQ) (Bruder et al., 2013). This scale consists of five statements, including “I think that politicians usually do not tell us the true motives for their decisions” and “I think that many very important things happen in the world, which the public is never informed about”. Participants rated their agreement on an 11-point Likert scale (0 = completely disagree, 10 = completely agree). The total CMQ score was calculated by summing responses across all items, with possible scores ranging from 0 to 50—higher scores reflecting stronger conspiracy beliefs. We used the validated Greek adaptation of the CMQ (Katsiroumpa, Moisoglou, Lamprakopoulou, et al., 2025), which demonstrated good internal consistency in our sample (Cronbach’s α = 0.845).

We used the Online Misinformation Susceptibility Scale (OMISS) (Katsiroumpa, Moisoglou, Mangoulia, et al., 2025) to measure online misinformation susceptibility in our sample. The OMISS includes nine items such as “When you see a post or story that interests you on social media or websites, how often do you check if the post includes the author’s name?” and “When you see a post or story that interests you on social media or websites, how often do you check if the post originates from a reliable source, such as authoritative news sites?”. Answers are on a five-point Likert scale; never (5), rarely (4), sometimes (3), very often (2), always (1). Total score ranges from 9 to 45. Higher scores indicate higher misinformation susceptibility. Developers of the OMISS suggest a cut-off point (≥23) (Katsiroumpa, Moisoglou, Gallos, et al., 2025) to distinguish individuals that show high levels of misinformation susceptibility from those who show normal levels of misinformation susceptibility. We used the valid Greek version of the OMISS (Katsiroumpa, Moisoglou, Mangoulia, et al., 2025). In our study, the Cronbach’s alpha for the OMISS was 0.927.

A widely used approach to assess individuals’ ability to detect fake news involves evaluating their responses to both true and false news headlines (Pennycook & Rand, 2019; Roozenbeek et al., 2020, 2022). In our study, participants were shown two fabricated and two factual headlines. To minimize national bias, particularly given that the study was conducted in Greece, all selected headlines excluded content related to Greek domestic affairs or national politics. Headlines were presented in plain text, without any visual elements or source indicators (such as specific websites or platforms). All items were current at the time of data collection. The two false headlines were sourced from the DiSiNFO Database (*DiSiNFO Database*, 2025), an initiative of the European Union’s Diplomatic Union (*Diplomatic Service of the European Union*, 2025) that involves experts in journalism, communication, and social sciences. The selected fake headlines were: (1) “US intelligence services read the messages of WhatsApp users” and (2) “Western mass media and social media are controlled by the government”. One true headline came from The Guardian (*Nasa Data Reveals Dramatic Rise in Intensity of Weather Events*, 2025), a respected British newspaper known for its authoritative and balanced journalism (*The Guardian*, 2025), and the other from France 24 (*France Mulls Social Media Ban for Under-15s after Fatal School Stabbing*, 2025), a Paris-based international news network funded by the French government (*France 24*, 2025). The selected true headlines were: (1) “NASA data reveals dramatic rise in intensity of weather events” and (2) “French President Emmanuel Macron wants to ban social media for under-15s in France”. Participants rated each headline by answering the question: “To the best of your knowledge, how likely is it that the claim in each headline is correct?”. Responses were given on a five-point Likert scale: (a) extremely unlikely, (b) somewhat unlikely, (c) neither unlikely nor likely, (d) somewhat likely, and (e) extremely likely. For fake headlines, scores were assigned as follows: 4 for “extremely unlikely,” 3 for “somewhat unlikely,” 2 for “neither,” 1 for “somewhat likely,” and 0 for “extremely likely”. For true headlines, the scoring was reversed. The scores for all four headlines were summed to produce a total score ranging from 0 to 16, with higher scores reflecting greater accuracy in detecting fake news.

### Ethical issues

Data were collected anonymously and voluntarily after participants were briefed on the study’s purpose and methodology and provided informed consent. The research adhered to the ethical guidelines outlined in the Declaration of Helsinki (“World Medical Association Declaration of Helsinki,” 2013). Additionally, the Ethics Committee of the Faculty of Nursing at the National and Kapodistrian University of Athens granted formal approval for the study protocol (approval No. 75, July 13, 2025).

## Statistical analysis

Categorical variables are reported as numbers and percentages, while continuous variables are presented as mean (SD) and median (interquartile range). Normality was assessed using the Kolmogorov-Smirnov and Q-Q plots, confirming that continuous variables followed a normal distribution. Independent variables included demographic factors, trust in scientists, and conspiracy beliefs, whereas dependent variables were online misinformation susceptibility and fake news detection. Given the normality of the dependent variables, we employed linear regression analysis. First, we conducted simple linear regression to assess univariate associations, followed by a multivariable regression model to determine the independent effects of each predictor. Results are reported as unadjusted and adjusted beta coefficients, 95% confidence intervals (CI), and p-values. Additionally, Pearson’s correlation coefficient was used to evaluate associations between normally distributed scale scores. A p-value < 0.05 was considered statistically significant. We used the IBM SPSS 28.0 (IBM Corp. Released 2021. IBM SPSS Statistics for Windows, Version 28.0. Armonk, NY: IBM Corp) for the analysis.

## Results

### Demographic characteristics

Table 1 presents the demographic details of the study participants. The sample comprised 342 individuals, with 71.3% being female. The mean age was 46.83 years (SD = 9.19), and the median age was 48 years (interquartile range = 12 years). The mean financial status score was 5.77 (SD = 1.46), with a median of 6 (interquartile range = 2). The mean trust score in websites was 3.70 (SD = 1.94), and the median score was 3 (interquartile range = 3). Interest in politics had a mean score of 5.89 (SD = 2.62) and a median of 6 (interquartile range = 3). Participants spent an average of 2.60 hours daily on the web or social media (SD = 2.19), with a median of 2 hours (interquartile range = 2 hours).

**Table 1.** Demographic characteristics of the study sample (n=342).

| Characteristics | N | % |
| --- | --- | --- |
| Gender |  |  |
| Males | 98 | 28.7 |
| Females | 244 | 71.3 |
| Age<sup>a</sup> | 46.83 | 9.19 |
| Educational level |  |  |
| High school | 46 | 13.5 |
| University | 115 | 33.6 |
| MSc/PhD diploma | 181 | 52.9 |
| Financial status<sup>a</sup> | 5.77 | 1.46 |
| Trust in websites<sup>a</sup> | 3.70 | 1.93 |
| Interest in politics<sup>a</sup> | 5.89 | 2.62 |
| Daily time in web/social media (hours)<sup>a</sup> | 2.60 | 2.19 |

<sup>a</sup> mean, standard deviation

### Study scales

Table 2 displays the descriptive statistics for the study scales. The mean score on the OMISS was 24.42 (SD = 8.85), with a median of 24. Additionally, 52.0% (n = 178) of participants showed high susceptibility to misinformation. The mean score for fake news detection was 8.36 (SD = 2.10), with a median of 8. The mean TISS score was 3.29 (SD = 0.68), while the CMQ had a mean score of 35.72 (SD = 9.47).

**Table 2.** Descriptive statistics for the study scales (n=342).

| Scale | Mean | Standard deviation | Median | Interquartile range |
| --- | --- | --- | --- | --- |
| Online Misinformation Susceptibility Scale | 24.42 | 8.85 | 24 | 12 |
| Fake news detection scale | 8.36 | 2.10 | 8 | 2 |
| Trust in Scientists Scale | 3.29 | 0.68 | 3.33 | 1 |
| Conspiracy Mentality Questionnaire | 35.72 | 9.47 | 36 | 13 |

Table 3 presents the correlations among the study scales. A negative correlation was observed between online misinformation susceptibility and both fake news detection (r =-0.141, p = 0.009) and trust in scientists (r =-0.417, p < 0.001). Additionally, online misinformation susceptibility showed a positive correlation with conspiracy beliefs (r = 0.188, p < 0.001). Fake news detection was positively correlated with trust in scientists (r = 0.144, p = 0.007) and negatively correlated with conspiracy beliefs (r =-0.329, p < 0.001). Lastly, there was a negative correlation between trust in scientists and conspiracy beliefs (r =-0.128, p = 0.018).

**Table 3.** Pearson’s correlation coefficients for the study scales (n=342).

| Scale | 2 | 3 | 4 |
| --- | --- | --- | --- |
| 1. Online Misinformation Susceptibility Scale | -0.141** | -0.417** | 0.181** |
| 2. Fake news detection scale |  | 0.144** | -0.329** |
| 3. Trust in Scientists Scale |  |  | -0.128* |
| 4. Conspiracy Mentality Questionnaire |  |  |  |

\* p-value < 0.05
\*\* p-value < 0.01

### Dependent variable: online misinformation susceptibility

Table 4 shows linear regression models with OMISS as the dependent variable. Our multivariable analysis identified trust in scientists and conspiracy behaviors as independent predictors of online misinformation susceptibility. In particular, we found that lower trust in scientists is associated with higher misinformation susceptibility (adjusted coefficient beta =-4.477, 95% CI =-5.752 to-3.203, p < 0.001). Participants who believed in conspiracy behaviors showed also higher levels of misinformation susceptibility (adjusted coefficient beta = 0.106, 95% CI = 0.015 to 0.196, p = 0.023). Our results showed that four demographic variables affect online misinformation susceptibility. In particular, lower financial status (adjusted coefficient beta =-0.948, 95% CI =-1.565 to-0.332, p = 0.003) and lower interest in politics (adjusted coefficient beta =-0.788, 95% CI =-1.128 to-0.448, p < 0.001) were associated with misinformation susceptibility. Additionally, we found a positive association between trust in websites and misinformation susceptibility (adjusted coefficient beta = 0.740, 95% CI = 0.281 to 1.198, p = 0.002). Finally, we found a negative association between age and misinformation susceptibility (adjusted coefficient beta =-0.116, 95% CI =-0.207 to-0.025, p = 0.012).

**Table 4.**
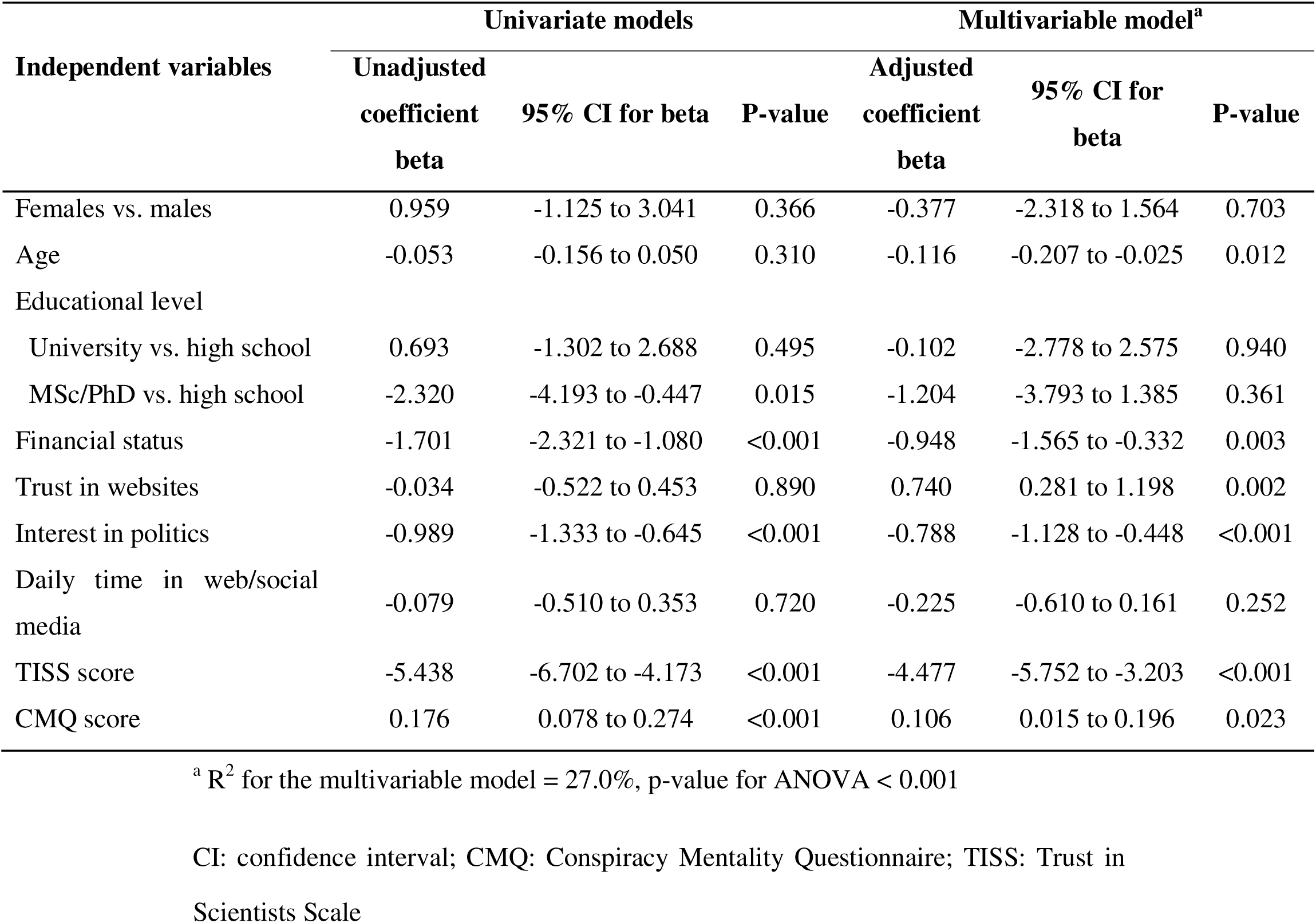
Linear regression models with Online Misinformation Susceptibility Scale as the dependent variable (n=342).

### Dependent variable: fake news detection

Table 5 shows the results from the linear regression analysis with fake news detection scale as the dependent variable. Multivariable linear regression analysis showed that participants who believe in conspiracy beliefs had a lower ability to detect fake news (adjusted coefficient beta =-0.051, 95% CI =-0.074 to-0.028, p < 0.001). Moreover, we found a positive association between fake news detection and financial status (adjusted coefficient beta = 0.189, 95% CI = 0.035 to 0.343, p = 0.016) and trust in sciences (adjusted coefficient beta = 0.235, 95% CI = 0.121 to 0.349, p < 0.001). Finally, we found a negative association between interest in politics and fake news detection (adjusted coefficient beta =-0.152, 95% CI =-0.237 to-0.068, p < 0.001).

**Table 5.** Linear regression models with fake news detection scale as the dependent variable (n=342).

| Independent variables | Univariate models |  |  | Multivariable model<sup>a</sup> |  |  |
| --- | --- | --- | --- | --- | --- | --- |
|  | Unadjusted coefficient beta | 95% CI for beta | P-value | Adjusted coefficient beta | 95% CI for beta | P-value |
| Females vs. males | -0.569 | -1.058 to -0.079 | 0.023 | -0.813 | -1.297 to -0.329 | 0.001 |
| Age | -0.017 | -0.041 to 0.007 | 0.167 | -0.011 | -0.034 to 0.011 | 0.325 |
| Educational level |  |  |  |  |  |  |
| University vs. high school | -0.568 | -1.037 to -0.100 | 0.018 | -0.342 | -1.009 to 0.325 | 0.314 |
| MSc/PhD vs. high school | 0.727 | 0.286 to 1.167 | 0.001 | 0.031 | -0.615 to 0.676 | 0.926 |
| Financial status | 0.312 | 0.163 to 0.461 | <0.001 | 0.189 | 0.035 to 0.343 | 0.016 |
| Trust in websites | 0.223 | 0.111 to 0.336 | <0.001 | 0.235 | 0.121 to 0.349 | <0.001 |
| Interest in politics | -0.047 | -0.132 to 0.038 | 0.275 | -0.152 | -0.237 to -0.068 | <0.001 |
| Daily time in web/social media | -0.040 | -0.142 to 0.062 | 0.436 | -0.080 | -0.176 to 0.017 | 0.104 |
| TISS score | 0.446 | 0.120 to 0.771 | 0.007 | 0.271 | -0.046 to 0.589 | 0.094 |
| CMQ score | -0.073 | -0.095 to -0.050 | <0.001 | -0.051 | -0.074 to -0.028 | <0.001 |

<sup>a</sup> $R^2$ for the multivariable model = 18.9%, $p$ -value for ANOVA < 0.001
CI: confidence interval; CMQ: Conspiracy Mentality Questionnaire; TISS: Trust in Scientists Scale

## Discussion

We found associations between trust in scientists, conspiracy beliefs, online misinformation susceptibility and fake news detection. Moreover, our findings revealed that several demographic variables affect online misinformation susceptibility and fake news detection.

Firstly, our results indicated that lower trust in scientists is associated with greater misinformation susceptibility and reduced ability to detect fake news. One plausible explanation for this association is rooted in the trust heuristic: individuals often rely on trusted sources as cognitive shortcuts when evaluating complex information. Reduced trust in scientists removes this heuristic, forcing individuals to rely on less credible or ideologically aligned sources, which frequently propagate misinformation. Consequently, diminished trust may not only increase exposure to false content but also impair critical evaluation skills, as skepticism toward legitimate expertise can spill over into generalized doubt about factual claims (Broda & Strömbäck, 2024; Lewandowsky et al., 2017; Lewandowsky & Van Der Linden, 2021). When individuals perceive scientists as unreliable or biased, they may disengage from evidence-based sources and become more vulnerable to alternative narratives, including misinformation. This pattern dovetails with converging evidence from COVID-19 and broader science-communication research showing that trust in scientific actors and institutions shapes information seeking, belief formation, and protective behaviors. For example, a two wave panel data during the pandemic in the United States suggest that distrust in science pushes people toward social media information sources, which in turn raises susceptibility to misperceptions (Lee et al., 2024). In similar fashion, a Czech longitudinal study found that trust in official information was the strongest predictor of vaccine acceptance, underscoring the practical significance of epistemic trust for real-world decision making (Grygarová et al., 2025). Recent evidence highlights that (i) misinformation about science is consequential and sticky, (ii) corrections are often only modestly effective, and (iii) trust and ideology moderate intervention success—factors that align with the gradients we observe by trust level. In the contemporary information ecosystem, even media attention to misinformation (not just exposure to misinformation itself) can erode trust in scientists—a dynamic that may compound susceptibility among low-trust audiences (Ecker et al., 2022; Hoes et al., 2025). Trust in scientists functions as a fast, ecologically rational cue for source reliability. When trust is low, people may (a) discount high-quality sources, (b) substitute social identity or partisan cues, and (c) increase reliance on social media intermediaries whose algorithms reward engagement over accuracy—together increasing vulnerability to false claims (Lee et al., 2024).

Additionally, our study demonstrates that individuals who endorse conspiracy beliefs exhibit greater susceptibility to misinformation and poorer ability to detect fake news. This finding aligns with prior research showing that conspiracy thinking is a strong predictor of epistemic vulnerability in digital environments (Douglas et al., 2017; Uscinski et al., 2022). Conspiratorial worldviews often involve distrust of authoritative sources, motivated reasoning, and confirmation bias, which collectively impair discernment between accurate and false information. Conspiracy beliefs typically involve assumptions of hidden agendas and malevolent actors, which can undermine confidence in authoritative sources and scientific consensus. Consequently, individuals may become more receptive to misinformation that aligns with their worldview and less capable of critically evaluating content. Conspiracy beliefs are rooted in cognitive biases such as proportionality bias and agency detection bias, which predispose individuals to accept elaborate, hidden explanations over mundane ones (Gagliardi, 2025). These biases interact with motivational factors—such as a desire for certainty and control—making conspiracy believers more receptive to misinformation narratives that validate their worldview (Zhou & Shen, 2024). A recent meta-analysis confirms that conspiracy ideation correlates strongly with intuitive thinking and low reflective reasoning, reducing the likelihood of engaging in deliberative processes that improve truth discernment (Yelbuz et al., 2022). For instance, during the COVID-19 pandemic, conspiracy beliefs were strongly associated with misinformation acceptance and vaccine hesitancy, illustrating the real-world consequences of this cognitive vulnerability (Bierwiaczonek et al., 2022). These patterns persist across domains, from health to politics, suggesting that conspiracy thinking is a generalized risk factor for misinformation susceptibility.

We found that lower financial status is associated with higher online misinformation susceptibility and lower levels of ability to detect fake news. Individuals with lower socioeconomic status often face structural disadvantages that limit their access to high-quality information and educational resources. These limitations can impair the development of critical thinking and digital literacy skills, which are essential for discerning credible information online. Sultan et al. found that analytical thinking significantly enhances the ability to discriminate between true and false news, a skill that tends to be more developed among individuals with higher education and financial stability (Sultan et al., 2024). Moreover, research demonstrated that people frequently overestimate their capacity to detect fake news, and those with lower socioeconomic status may lack the opportunities for feedback and learning that foster accurate self-assessment and improvement (Assenza et al., 2024). Economic hardship can also increase psychological stress, which has been linked to greater receptivity to emotionally charged misinformation (Roozenbeek et al., 2023). Additionally, individuals with lower financial status may rely more heavily on social media for news due to cost barriers, exposing them to higher volumes of unverified or misleading content (Broda & Strömbäck, 2024). These factors collectively contribute to a heightened vulnerability to misinformation and a diminished capacity for fake news detection among economically disadvantaged populations.

In this study, individuals exhibiting lower levels of political interest were more prone to accepting online misinformation and demonstrated diminished proficiency in discerning fake news. This association can be understood through the role of political engagement in fostering critical evaluation and media literacy skills. Individuals with limited political interest often consume fewer credible news sources and lack exposure to fact-checking practices, which reduces their ability to discern accurate information from falsehoods (Bowles et al., 2025). Political disengagement is also linked to lower motivation to verify information and a reliance on heuristic processing, making emotionally charged or sensational content more persuasive (Pantazi et al., 2021). Furthermore, political knowledge and critical thinking have been identified as protective factors against misinformation, as they encourage skepticism toward unverified claims and promote analytical reasoning (List et al., 2024). Research on misinformation receptivity frameworks highlights that individuals with weak prior knowledge are more likely to accept misleading narratives, particularly in noisy communication environments (Zmigrod et al., 2023). Additionally, a recent systematic review indicates that political misinformation thrives in contexts of low engagement, where individuals lack the cognitive and informational resources to challenge deceptive narratives (Broda & Strömbäck, 2024). Collectively, these findings underscore the importance of political interest as a cognitive and motivational buffer against misinformation vulnerability.

The results of this study indicate an inverse association between age and susceptibility to misinformation, with younger participants exhibiting a higher propensity to accept misleading information. This trend can be attributed to several factors. Younger individuals typically engage more frequently with social media platforms, which serve as primary channels for misinformation dissemination, thereby increasing exposure to misleading content. Despite being digitally adept, younger cohorts often lack advanced media literacy and critical evaluation skills, making them less capable of distinguishing credible sources from false narratives (GherguC-Babii et al., 2025).

Furthermore, research suggests that younger adults rely more on heuristic processing and exhibit stronger motivations to share information online, which amplifies their susceptibility to misinformation (Jo et al., 2024). In contrast, older adults benefit from higher levels of crystallized intelligence and accumulated knowledge, which enhance their ability to detect fake news despite slower cognitive processing (Golino et al., 2023). Studies confirm that Generation Z and Millennials score lower on misinformation detection tests compared to older age groups, challenging assumptions that digital natives are inherently better at navigating online information environments (Kyrychenko et al., 2025).

## Limitations

Our study had several limitations warrant consideration. First, the cross-sectional design limits causal inference; while low trust in scientists and conspiracy beliefs predicted misinformation vulnerability, reverse causality is plausible—exposure to misinformation may erode trust in scientists over time. In other words, it is plausible that exposure to misinformation reinforces conspiratorial thinking and distrust in scientists. Therefore, longitudinal studies are necessary to further investigate the association between the study variables. Second, the study relied on self-reported measures, which may be subject to social desirability bias. Although we used valid scales to measure our study variables, information bias is still probable. Third, we employed a convenience sample of the population of a specific country. Thus, our findings cannot be generalized even to the broader population of this country. Moreover, beliefs about science, conspiracy theories and misinformation differ across cultures, so findings may be context-specific. Cultural and ideological contexts influence both trust and conspiracy prevalence and misinformation vulnerability. Therefore, scholars should conduct further studies in different countries with different cultural contexts to obtain more valid information. Additionally, random or/and stratified samples of the general population should be recruited to obtain results than can be generalized. Fourth, we investigated the influence of several demographic variables on misinformation susceptibility and fake news detection. However, several other factors could be investigated as potential predictors such as political ideology and media literacy. Fifth, trust and conspiracy beliefs may interact with other psychological traits (e.g. personality traits, cognitive style, fear, anxiety) that could be measured in future studies to infer more valid results. Finally, online misinformation evolves rapidly, so stimuli used in the study might become outdated a few months later. Thus, there is a need to measure levels of online misinformation susceptibility through time by employing longitudinal studies.

## Conclusions

The implications of our findings are significant for public health communication, science education, and policy-making. Strengthening trust in scientists should be a priority for combating misinformation. Strategies may include (a) transparent communication about scientific processes and uncertainties, (b) active engagement with communities to address concerns and misconceptions, and (c) media literacy programs that integrate understanding of scientific rigor and peer review mechanisms. Such interventions could reduce misinformation susceptibility by reinforcing confidence in credible sources and improving critical evaluation skills.

Additionally, efforts to reduce misinformation vulnerability should consider the role of conspiracy beliefs as a risk factor. In this case, policy makers should improve community-based engagement to rebuild trust in credible sources without dismissing concerns outright. Also, critical thinking and media literacy programs tailored to address cognitive biases associated with conspiracy thinking could reduce misinformation susceptibility and increase individuals’ ability to detect fake news. Moreover, interventions based on inoculation theory—such as exposing individuals to weakened forms of misinformation and explaining manipulation tactics—have shown promise in reducing susceptibility (Van Der Linden et al., 2017). These approaches could help mitigate the influence of conspiracy beliefs on information processing and improve resilience against misinformation.

Future research should employ longitudinal designs to clarify causal pathways between trust in science, conspiracy beliefs, misinformation susceptibility and fake news detection. Moreover, investigation of moderating/mediating variables, such as political ideology, media consumption patterns, and cognitive biases could add significant information. Additionally, scholars should develop intervention programs aimed at rebuilding trust and enhancing critical thinking skills, and also explore their effectiveness. Understanding these mechanisms will inform targeted strategies to reduce misinformation vulnerability and promote evidence-based decision-making in increasingly complex information ecosystems.

## Data Availability

All data produced are available online at
https://doi.org/10.6084/m9.figshare.30646349

https://doi.org/10.6084/m9.figshare.30646349

## References

Aïmeur, E., Amri, S., & Brassard, G. (2023). Fake news, disinformation and misinformation in social media: A review. Social Network Analysis and Mining, 13(1), 30. 10.1007/s13278-023-01028-5

Ali Adeeb, R., & Mirhoseini, M. (2023). The Impact of Affect on the Perception of Fake News on Social Media: A Systematic Review. Social Sciences, 12(12), 674. 10.3390/socsci12120674

Assenza, T., Cardaci, A., & Huber, S. (2024). Fake news: Susceptibility, awareness and solutions (TSE Working Paper). Toulouse School of Economics.

Bago, B., Rand, D. G., & Pennycook, G. (2020). Fake news, fast and slow: Deliberation reduces belief in false (but not true) news headlines. Journal of Experimental Psychology: General, 149(8), 1608–1613. 10.1037/xge0000729

Bierwiaczonek, K., Gundersen, A. B., & Kunst, J. R. (2022). The role of conspiracy beliefs for COVID-19 health responses: A meta-analysis. Current Opinion in Psychology, 46, 101346. 10.1016/j.copsyc.2022.101346

Borukhson, D., Lorenz-Spreen, P., & Ragni, M. (2022). When Does an Individual Accept Misinformation? An Extended Investigation Through Cognitive Modeling. Computational Brain & Behavior, 5(2), 244–260. 10.1007/s42113-022-00136-3

Bowles, J., Croke, K., Larreguy, H., Liu, S., & Marshall, J. (2025). Sustaining Exposure to Fact-Checks: Misinformation Discernment, Media Consumption, and Its Political Implications. American Political Science Review, 119(4), 1864–1887. 10.1017/S0003055424001394

Broda, E., & Strömbäck, J. (2024). Misinformation, disinformation, and fake news: Lessons from an interdisciplinary, systematic literature review. Annals of the International Communication Association, 48(2), 139–166. 10.1080/23808985.2024.2323736

Bruder, M., Haffke, P., Neave, N., Nouripanah, N., & Imhoff, R. (2013). Measuring Individual Differences in Generic Beliefs in Conspiracy Theories Across Cultures: Conspiracy Mentality Questionnaire. Frontiers in Psychology, 4, 1–15. 10.3389/fpsyg.2013.00225

Butter, M., & Knight, P. (Eds.). (2020). Routledge Handbook of Conspiracy Theories (1st ed.). Routledge. 10.4324/9780429452734

Cologna, V., Mede, N. G., Berger, S., Besley, J., Brick, C., Joubert, M., Maibach, E. W., Mihelj, S., Oreskes, N., Schäfer, M. S., Van Der Linden, S., Abdul Aziz, N. I., Abdulsalam, S., Shamsi, N. A., Aczel, B., Adinugroho, I., Alabrese, E., Aldoh, A., Alfano, M.,…Zwaan, R. A. (2025). Trust in scientists and their role in society across 68 countries. Nature Human Behaviour, 9(4), 713–730. 10.1038/s41562-024-02090-5

Del Vicario, M., Bessi, A., Zollo, F., Petroni, F., Scala, A., Caldarelli, G., Stanley, H. E., & Quattrociocchi, W. (2016). The spreading of misinformation online. Proceedings of the National Academy of Sciences, 113(3), 554–559. 10.1073/pnas.1517441113

*Diplomatic Service of the European Union*. (2025, June 17). Diplomatic Service of the European Union. https://www.eeas.europa.eu/_en

*DiSiNFO Database*. (2025, June 17). EU vs DiSiNFO. https://euvsdisinfo.eu/disinformation-cases/

Douglas, K. M., Sutton, R. M., & Cichocka, A. (2017). The Psychology of Conspiracy Theories. Current Directions in Psychological Science, 26(6), 538–542. 10.1177/0963721417718261

Ecker, U. K. H., Lewandowsky, S., & Chadwick, M. (2020). Can corrections spread misinformation to new audiences? Testing for the elusive familiarity backfire effect. Cognitive Research: Principles and Implications, 5(1), 41. 10.1186/s41235-020-00241-6

Ecker, U. K. H., Lewandowsky, S., Cook, J., Schmid, P., Fazio, L. K., Brashier, N., Kendeou, P., Vraga, E. K., & Amazeen, M. A. (2022). The psychological drivers of misinformation belief and its resistance to correction. Nature Reviews Psychology, 1(1), 13–29. 10.1038/s44159-021-00006-y

Erisen, C., & Erisen, E. (2024). Populist Attitudes and Misinformation Challenging Trust: The Case of Turkey. International Journal of Public Opinion Research, 37(1), edae056. 10.1093/ijpor/edae056

Forgas, J. P. (2025). The psychology of false beliefs: Collective delusions and conspiracy theories. Routledge.

*France 24*. (2025, June 17). Wikipedia. https://en.wikipedia.org/wiki/France_24

*France mulls social media ban for under-15s after fatal school stabbing*. (2025, June 17). France24. https://www.france24.com/en/live-news/20250611-france-eyes-social-media-ban-for-under-15s-after-school-stabbing

Freeman, D., & Bentall, R. P. (2017). The concomitants of conspiracy concerns. Social Psychiatry and Psychiatric Epidemiology, 52(5), 595–604. 10.1007/s00127-017-1354-4

Gagliardi, L. (2025). The role of cognitive biases in conspiracy beliefs: A literature review. Journal of Economic Surveys, 39(1), 32–65. 10.1111/joes.12604

GherguC-Babii, A.-N., Poleac, G., & Obadă, D.-R. (2025). Challenges for NGO Communication Practitioners in the Disinformation Era: A Qualitative Study Exploring Generation Z’s Perception of Civic Engagement and Their Vulnerability to Online Fake News. Journalism and Media, 6(3), 136. 10.3390/journalmedia6030136

Green, R., Toribio-Flórez, D., & Douglas, K. M. (2023). Impressions of science and healthcare professionals who share anti-science conspiracy theories. Routledge Open Research, 2, 37. 10.12688/routledgeopenres.17965.1

Grygarová, D., Kožený, J., Tišanská, L., Havlík, M., & Horáček, J. (2025). Trust in official information as a key predictor of COVID-19 vaccine acceptance: Evidence from a Czech longitudinal survey study. BMC Public Health, 25(1), 770. 10.1186/s12889-025-21988-x

Hameleers, M., & Brosius, A. (2022). You Are Wrong Because I Am Right! The Perceived Causes and Ideological Biases of Misinformation Beliefs. International Journal of Public Opinion Research, 34(1), edab028. 10.1093/ijpor/edab028

Hatton, C. R., Barry, C. L., Levine, A. S., McGinty, E. E., & Han, H. (2022). American Trust in Science & Institutions in the Time of COVID-19. Daedalus, 151(4), 83–97. 10.1162/daed_a_01945

Hoes, E., Clemm, B., Gessler, T., Qian, S., & Wojcieszak, M. (2025). (Media Attention to) Misinformation Can Undermine Trust in Scientists. Political Behavior. 10.1007/s11109-025-10090-y

Hubeny, T. J., Nahon, L. S., Ng, N. L., & Gawronski, B. (2025). Who Falls for Misinformation and Why? Personality and Social Psychology Bulletin, 01461672251328800. 10.1177/01461672251328800

Jo, H., Yang, F., & Yan, Q. (2024). Spreaders vs victims: The nuanced relationship between age and misinformation via FoMO and digital literacy in different cultures. New Media & Society, 26(9), 5169–5194. 10.1177/14614448221130476

Jolley, D., & Douglas, K. M. (2014). The social consequences of conspiracism: Exposure to conspiracy theories decreases intentions to engage in politics and to reduce one’s carbon footprint. British Journal of Psychology, 105(1), 35–56. 10.1111/bjop.12018

Katsiroumpa, A., Moisoglou, I., Gallos, P., Galani, O., Tsiachri, M., Lamprakopoulou, K., & Galanis, P. (2025). Online Misinformation Susceptibility Scale: Determination of an optimal cut-off point. *International Journal of Caring Sciences*, Under press.

Katsiroumpa, A., Moisoglou, I., Lamprakopoulou, K., Galani, O., Tsiachri, M., Konstantakopoulou, O., & Galanis, P. (2025). Conspiracy Mentality Questionnaire: Translation and validation in Greek. *International Journal of Caring Sciences*, Under press.

Katsiroumpa, A., Moisoglou, I., Mangoulia, P., Konstantakopoulou, O., Gallos, P., Tsiachri, M., & Galanis, P. (2025). The Online Misinformation Susceptibility Scale: Development and Initial Validation. Healthcare, 13(17), 2252. 10.3390/healthcare13172252

Kaufman, R., Broukhim, A., & Haupt, M. (2025). WARNING This Contains Misinformation: The Effect of Cognitive Factors, Beliefs, and Personality on Misinformation Warning Tag Attitudes. Proceedings of the ACM on Human-Computer Interaction, 9(7), 1–32. 10.1145/3757521

Kim, S., Capasso, A., Ali, S. H., Headley, T., DiClemente, R. J., & Tozan, Y. (2022). What predicts people’s belief in COVID-19 misinformation? A retrospective study using a nationwide online survey among adults residing in the United States. BMC Public Health, 22(1), 2114. 10.1186/s12889-022-14431-y

Kyrychenko, Y., Koo, H. J., Maertens, R., Roozenbeek, J., Van Der Linden, S., & Götz, F. M. (2025). Profiling misinformation susceptibility. Personality and Individual Differences, 241, 113177. 10.1016/j.paid.2025.113177

Lazer, D. M. J., Baum, M. A., Benkler, Y., Berinsky, A. J., Greenhill, K. M., Menczer, F., Metzger, M. J., Nyhan, B., Pennycook, G., Rothschild, D., Schudson, M., Sloman, S. A., Sunstein, C. R., Thorson, E. A., Watts, D. J., & Zittrain, J. L. (2018). The science of fake news. Science, 359(6380), 1094–1096. 10.1126/science.aao2998

Lee, S., Jones-Jang, S. M., Chung, M., Lee, E. W. J., & Diehl, T. (2024). Examining the Role of Distrust in Science and Social Media Use: Effects on Susceptibility to COVID Misperceptions with Panel Data. Mass Communication and Society, 27(4), 653–678. 10.1080/15205436.2023.2268053

Lewandowsky, S., Ecker, U. K. H., & Cook, J. (2017). Beyond misinformation: Understanding and coping with the “post-truth” era. Journal of Applied Research in Memory and Cognition, 6(4), 353–369. 10.1016/j.jarmac.2017.07.008

Lewandowsky, S., & Van Der Linden, S. (2021). Countering Misinformation and Fake News Through Inoculation and Prebunking. European Review of Social Psychology, 32(2), 348–384. 10.1080/10463283.2021.1876983

List, J. A., Ramirez, L. M., Seither, J., Unda, J., & Vallejo, B. H. (2024). Critical thinking and misinformation vulnerability: Experimental evidence from Colombia. PNAS Nexus, 3(10), pgae361. 10.1093/pnasnexus/pgae361

*Nasa data reveals dramatic rise in intensity of weather events*. (2025, June 17). The Guardian. https://www.theguardian.com/world/2025/jun/17/nasa-data-reveals-dramatic-rise-in-intensity-of-weather-events

O’Brien, T. C., Palmer, R., & Albarracin, D. (2021). Misplaced trust: When trust in science fosters belief in pseudoscience and the benefits of critical evaluation. Journal of Experimental Social Psychology, 96, 104184. 10.1016/j.jesp.2021.104184

Pantazi, M., Hale, S., & Klein, O. (2021). Social and Cognitive Aspects of the Vulnerability to Political Misinformation. Political Psychology, 42(S1), 267–304. 10.1111/pops.12797

Pennycook, G., & Rand, D. G. (2019). Lazy, not biased: Susceptibility to partisan fake news is better explained by lack of reasoning than by motivated reasoning. Cognition, 188, 39–50. 10.1016/j.cognition.2018.06.011

Pennycook, G., & Rand, D. G. (2021). The Psychology of Fake News. Trends in Cognitive Sciences, 25(5), 388–402. 10.1016/j.tics.2021.02.007

Piksa, M., Noworyta, K., Piasecki, J., Gwiazdzinski, P., Gundersen, A. B., Kunst, J., & Rygula, R. (2022). Cognitive Processes and Personality Traits Underlying Four Phenotypes of Susceptibility to (Mis)Information. Frontiers in Psychiatry, 13, 912397. 10.3389/fpsyt.2022.912397

Roozenbeek, J., Culloty, E., & Suiter, J. (2023). Countering Misinformation: Evidence, Knowledge Gaps, and Implications of Current Interventions. European Psychologist, 28(3), 189–205. 10.1027/1016-9040/a000492

Roozenbeek, J., Maertens, R., Herzog, S. M., Geers, M., Kurvers, R., Sultan, M., & Van Der Linden, S. (2022). Susceptibility to misinformation is consistent across question framings and response modes and better explained by myside bias and partisanship than analytical thinking. Judgment and Decision Making, 17(3), 547–573. 10.1017/S1930297500003570

Roozenbeek, J., Schneider, C. R., Dryhurst, S., Kerr, J., Freeman, A. L. J., Recchia, G., Van Der Bles, A. M., & Van Der Linden, S. (2020). Susceptibility to misinformation about COVID-19 around the world. Royal Society Open Science, 7(10), 201199. 10.1098/rsos.201199

Scherer, L. D., & Pennycook, G. (2020). Who Is Susceptible to Online Health Misinformation? American Journal of Public Health, 110(S3), S276–S277. 10.2105/AJPH.2020.305908

Sultan, M., Tump, A. N., Ehmann, N., Lorenz-Spreen, P., Hertwig, R., Gollwitzer, A., & Kurvers, R. H. J. M. (2024). Susceptibility to online misinformation: A systematic meta-analysis of demographic and psychological factors. Proceedings of the National Academy of Sciences, 121(47), e2409329121. 10.1073/pnas.2409329121

*The Guardian*. (2025, June 17). Wikipedia. https://en.wikipedia.org/wiki/The_Guardian

Toribio-Flórez, D., Green, R., Sutton, R. M., & Douglas, K. M. (2023). Does Belief in Conspiracy Theories Affect Interpersonal Relationships? The Spanish Journal of Psychology, 26, e9. 10.1017/SJP.2023.8

Udry, J., & Barber, S. J. (2024). The illusory truth effect: A review of how repetition increases belief in misinformation. Current Opinion in Psychology, 56, 101736. 10.1016/j.copsyc.2023.101736

Uscinski, J., Enders, A., Klofstad, C., Seelig, M., Drochon, H., Premaratne, K., & Murthi, M. (2022). Have beliefs in conspiracy theories increased over time? PLOS ONE, 17(7), e0270429. 10.1371/journal.pone.0270429

Van Der Linden, S., Leiserowitz, A., Rosenthal, S., & Maibach, E. (2017). Inoculating the Public against Misinformation about Climate Change. Global Challenges, 1(2), 1600008. 10.1002/gch2.201600008

Von Elm, E., Altman, D. G., Egger, M., Pocock, S. J., Gøtzsche, P. C., & Vandenbroucke, J. P. (2008). The Strengthening the Reporting of Observational Studies in Epidemiology (STROBE) statement: Guidelines for reporting observational studies. Journal of Clinical Epidemiology, 61(4), 344–349. 10.1016/j.jclinepi.2007.11.008

World Medical Association Declaration of Helsinki: Ethical Principles for Medical Research Involving Human Subjects. (2013). JAMA, 310(20), 2191. 10.1001/jama.2013.281053

Yelbuz, B. E., Madan, E., & Alper, S. (2022). Reflective thinking predicts lower conspiracy beliefs: A meta-analysis. Judgment and Decision Making, 17(4), 720–744. 10.1017/S1930297500008913

Zhou, Y., & Shen, L. (2024). Processing of misinformation as motivational and cognitive biases. Frontiers in Psychology, 15, 1430953. 10.3389/fpsyg.2024.1430953

Zmigrod, L., Burnell, R., & Hameleers, M. (2023). The Misinformation Receptivity Framework: Political Misinformation and Disinformation as Cognitive Bayesian Inference Problems. European Psychologist, 28(3), 173–188. 10.1027/1016-9040/a000498

